# The Association between Active Membership in Voluntary Organizations and Homonegativity

**DOI:** 10.1101/2024.03.07.24303952

**Authors:** Amirhossein Khanehpaza, Krit Pongpirul, Atthanee Jeeyapant

**Affiliations:** Health Development Program, Faculty of Medicine, Chulalongkorn University. Email: |; Associate Professor, Faculty of Medicine, Chulalongkorn University. Adjunct Faculty, Johns Hopkins Bloomberg School of Public Health. Research & Quality Counselor, Bumrungrad International Hospital.; Biostatistics Excellence Center, Research Affairs, Faculty of Medicine, Chulalongkorn University., Mahidol Oxford Tropical Medicine Research Unit, Faculty of Tropical Medicine, Mahidol University.

## Abstract

Homonegativity adversely affects the health and well-being of homosexuals in society, making it vital to identify factors associated with it. This study investigates whether active membership in voluntary organizations correlates with homonegativity. We analyzed the latest World Values Survey dataset, spanning 2017 to 2022, which encompasses 63 countries. Our findings suggest that active membership in certain voluntary organizations correlates with homonegativity levels among both men and women across various age groups. Specifically, active participation in sports or recreational organizations, professional associations, art, music, or educational organizations, and humanitarian or charitable organizations was found to be negatively correlated with homonegativity in specific gender-age groups, albeit with varying degrees of association strength. We postulate that this correlation might hint at causation, suggesting that active membership in these organizations promotes greater tolerance towards homosexuals and homosexuality. Furthermore, our analysis indicates that demographic and socio-economic variables, the political freedom of the respondent’s country, and the respondent’s life satisfaction are also linked to homonegativity.

## Introduction

The term “homonegativity” refers to negative reactions or attitudes towards homosexuality (Rye & Meaney, 2010; Moreno et al., 2015). Assessing the levels and determinants of homonegativity within societies is crucial, as numerous studies have indicated its severe impact on the health and well-being of homosexuals. For instance, homosexuals who face homonegativity often grapple with feelings of shame and self-loathing, leading to isolation (Thepsourinthone et al., 2021), depression, anxiety, poor mental health (Plöderl & Tremblay, 2015), as well as alcohol abuse and even suicide (Michli & Jamil, 2022). Given the profound consequences of homonegativity on the wellbeing of homosexuals, it is imperative to understand its determinants. Past research has highlighted several factors that influence homonegativity, including religion, ethnicity, gender, education level, economic development, and more (Meladze & Brown, 2015; Doebler, 2015; Watt & Elliot, 2019; Stulhofer & Rimac, 2009; Marsh & Brown, 2011). In this study, our objective is to determine whether active membership in voluntary organizations is linked to homonegativity.

## Literature review

Volunteerism is defined as the engagement in activities without the expectation of financial compensation. The existing research exploring the relationship between volunteerism and homonegativity is relatively limited. A significant study highlighted the connection between homophobia and voluntary participation in extracurricular activities among high school students. The results suggested that male students who engaged in mainstream sports (e.g., football, basketball) were more likely to exhibit negative attitudes towards homosexuality compared to their counterparts who didn’t participate in such sports. Conversely, female students who took part in non-athletic extracurricular activities, like debate and science clubs, were less likely to harbor homonegative sentiments compared to those who didn’t engage in these activities. These findings underscore the influence of both gender and the nature of voluntary activity in shaping the correlation between volunteering and attitudes towards homosexuality.

## Research questions

1. Can participation in voluntary activities predict homonegativity?
2. Does the relationship between voluntary activities and homonegativity vary based on the type of activity?
3. Does gender influence the relationship between voluntary activities and homonegativity?
4. Is the relationship between voluntary activities and homonegativity consistent across all age groups?

## Methodology

For this study, we utilized secondary data. The specifics of our methodology are detailed below.

### Data Source

The World Values Survey (WVS) stands as the most expansive non-commercial academic social survey program. Since its inception in 1981, it has conducted surveys across over 120 countries, boasting over 60,000 citations in Google Scholar alone (WVS Database, n.d.). WVS provides freely downloadable data sets for various countries on their website. We secured permission to use their data through email correspondence. Our analysis centers on the latest World Values Survey dataset (Wave 7), gathered between 2017 and 2022, which encompasses data from 63 countries (Haerpfer, C., Inglehart et al., 2022).

### Analytical Approach

After downloading the dataset in SPSS format, we employed binary logistic regression for data analysis. This analysis was executed using the SPSS version 28 software.

## Research Design

Our research designis descriptive in nature. We employed binary logistic regression for each age group to ascertain the association between explanatory variables and homonegativity.

## Variable Measurement

### Dependent Variable

In the WVS survey, respondents were posed the following question: “Please tell me for each of the following actions whether you think it can always be justified, never be justified, or something in between.” Among the actions listed was “homosexuality.” Respondents were prompted to select a number between 1 and 10, with 1 representing “never justified” and 10 indicating “always justified.” For the purposes of our study, we transformed this variable into a binary format. Responses ranging from 1-5 were categorized as ‘Homonegative’, while those between 6-10 were classified as ‘Not homonegative’. Consequently, our dependent variable is labeled ‘homonegative’, with binary outcomes of ‘Yes’ and ‘No’. Notably, 69.2% of the survey respondents fell into the ‘homonegative’ category.

## Explanatory Variables

### Our study incorporates a variety of explanatory variables

Socioeconomic Variables: These include factors such as income, education, employment status, and life satisfaction.

Demographic Variables: This set comprises age, gender, marital status, religiosity, and area of residence.

Volunteering Variables: Separate from the demographic factors, these variables specifically address the nature and extent of respondents’ volunteering activities.

However, to directly address our research questions, we have excluded age and gender from the general set of explanatory variables. Instead, we categorized respondents into three distinct age brackets and two gender classifications, culminating in a total of six combined age-gender groups. Table 1 provides a detailed breakdown of respondents across these combinations.

**Table 1:** Sample Size for Each Gender-Age Group.

| Age | Gender | Respondents |
| --- | --- | --- |
| 16-29 (21,753) | Male | 10,251 |
|  | Female | 11,502 |
| 30-49 (34,839) | Male | 16,192 |
|  | Female | 18,647 |
| 50+ (31,185) | Male | 15,115 |
|  | Female | 16,070 |

Below, we detail how our socioeconomic and demographic explanatory variables were derived from the survey:

1. Marital Status: We transformed this into a binary variable with outcomes of “Married” (or living together as if married) and “Otherwise”.
2. Religiosity: Respondents were asked: “Independently of whether you attend religious services or not, would you say you are a religious person, not a religious person or an atheist?” We classified those who chose the first option as ‘Religious’ and the latter two options as ‘Not Religious’.
3. Employment: Respondents were simply queried if they had ‘Paid Employment’ or not.
4. Education: We categorized those with a university degree as having a “High Level of Education”. Those with education below a university level were labeled as having a “Low Level of Education”.
5. Income: Respondents rated their income on a scale, with 1 indicating the lowest income group and 10 the highest in their country, accounting for all sources of income. We transformed this into a binary outcome: responses from 1 to 5 were classified as ‘Low Income’, while those from 6 to 10 were classified as ‘High Income’.
6. Life Satisfaction: Respondents were asked: “All things considered, how satisfied are you with your life as a whole these days?” with a choice scale from 1 (completely dissatisfied) to 10 (completely satisfied). This was converted into a binary variable: responses from 1-5 were labeled ‘Dissatisfied’, and those from 6-10 as ‘Satisfied’.
7. Area of Living: This binary variable indicates whether the respondent resides in an Urban or Rural area.

In addition to the previously introduced explanatory variables— income, education, employment, marital status, religiosity, life satisfaction, and area of living— we also factored in two more variables. While these were only included in select models (the rationale for which will be detailed in the ‘data analysis section’), they bear significance to our study.

The additional explanatory variables are:

1. Children: This binary variable indicates whether the respondent has any children.
2. Freedom of Political Regime: Among the myriad of variables contained within the WVS data, one stands out— the “type of political regime,” derived from the 2019 ‘Freedom House’ data (Countries and Territories, n.d.). This variable delineates the political freedom of the respondent’s country of residence, categorized into three tiers: free, partly free, and not free. For the purpose of our study, we bifurcated this into two broader categories. Countries labeled as “free” retained the designation ‘Free’ (comprising 37.6% of the data), while those marked as “partly free” or “not free” were grouped under ‘Not Free’. The inclusion of this variable is driven by discernible variances in average homonegativity levels across different nations (Jäckle & Wenzelburger, 2015). By integrating this variable, our goal is to ascertain the extent to which a country’s political freedom predicts the homonegative sentiments of its inhabitants.

We utilized binary logistic regression for data analysis and model generation. When integrating categorical variables into logistic regression, it’s essential to designate a reference category and formulate dummy variables for each of the remaining categories, setting the dummy variable to 0 for the reference category.

## Explanatory Variables Regarding ‘Voluntary Activity’

In the WVS survey, respondents were prompted about their membership status in various voluntary organizations. The question posed was:

“Now I am going to read off a list of voluntary organizations. For each organization, could you tell me whether you are an active member, an inactive member, or not a member of that type of organization?”

Respondents had three choices for each of the 12 organizations: ‘Not a member’, ‘Inactive member’, and ‘Active member’. We aimed to convert these options into binary variables, marking a dummy as 1 for ‘Active member’ and 0 for both ‘Inactive member’ and ‘Not a member’. The rationale behind this conversion stems from the ambiguity of what ‘Inactive membership’ truly signifies. Does it merely denote a registered but non-participatory member or someone who occasionally engages? Due to this lack of clarity, we structured our binary variable as ‘Active membership’ vs. all others.

Furthermore, we introduced an additional variable, termed ‘V-number’, that quantifies the count of voluntary organizations where the respondent is an active member. This variable is categorical, with outcomes being:

0 organizations (57% of respondents)
1 organization (21.1% of respondents)
More than 1 organization (21.9% of respondents)

The creation of ‘V-number’ aims to investigate if varying degrees of active volunteerism correlate with homonegativity. In this structure, the reference category is set as 0 organizations.

## Data Analysis

Binary logistic regression is employed when the dependent variable is binary. In our study, this binary variable is ‘homonegativity.’

## Models Employed for Analysis

For each age group, we formulated two models. Here, we detail the construction of Model A: To establish Model A, we followed these steps:

Each of the 9 primary explanatory variables, along with the V-number variable, was individually introduced into a univariate logistic regression. The dependent variable for these regressions was ‘homonegativity.’

From these individual regressions, only those explanatory variables that exhibited statistical significance were shortlisted.

These shortlisted variables were then combined in a multivariate binary logistic regression to formulate Model A.

While constructing this model, we also evaluated the potential inclusion of variables E8 (children) and E9 (political freedom). Their incorporation was based on ensuring they did not compromise the model’s fit to the data. For example, if introducing E8 led the Hosmer-Lemeshow test’s significance to fall below 0.05, we would omit E8, regardless of its statistical significance in the preceding univariate logistic regression.

## Model B

The primary aim of Model B is to discern the association between specific types of voluntary organizations and homonegativity.

### Selection Criteria

For the construction of this model, we consider only those respondents who are active members of either zero or just one organization. This decision is grounded in the rationale that for respondents with multiple memberships, pinpointing the precise organization influencing their stance on homonegativity becomes challenging. Notably, 57% of respondents were not active in any voluntary organization, while 21.1% were actively engaged in just one. Model B specifically includes these subsets of respondents.

### Construction Process

Similar to Model A, the creation of Model B commences by individually inserting the explanatory variables (including E1 to E9 and V1 to V12) into a univariate logistic regression.

Variables that exhibit statistical significance are subsequently incorporated into a multivariate logistic regression. Notably, the V-number variable is omitted in Model B.

Within this multivariate analysis, any explanatory variable that doesn’t demonstrate statistical significance—possibly due to a confounding effect—is methodically excluded. This pruning continues until all the remaining variables show statistical significance (P<0.05).

### Results Presentation

The results section will encompass the Omnibus test, Nagelkerke R-squared value, Hosmer-Lemeshow test, and the area under the Receiver Operating Characteristic (ROC) curve for both Models A and B across each of the six gender-age groups—culminating in a potential total of 12 models.

Both Model A and Model B will be presented distinctly, accompanied by the relative coefficient, Wald value, and odds ratio (along with its 95% confidence interval) for all retained explanatory variables.

## Handling Missing Data

When utilizing secondary data, the issue of missing data becomes critically important. Throughout our analysis, instances with missing data were excluded.

### Threshold for Missing Data

A common threshold in research suggests that if missing data accounts for less than 20% of the dataset, it’s generally considered acceptable (School of Transportation Engineering, Suranaree University of Technology & Meeyai, 2016).

### Our Data Analysis

Group B Models: In our Group B models, the percentage of missing data consistently fell below the 20% threshold (averaging 16.3% with a standard deviation of 2.7%). However, one exception was observed for women in age group 3, where the missing data accounted for 20.8%.

Group A Models: For the men and women in age groups 2 and 3 (comprising four models), the missing data ranged from 22% to 29%.

Given the slightly elevated missing data percentages in some models, we felt it prudent to conduct a sensitivity analysis.

### Sensitivity Analysis

For each model, we randomly excluded 25% of the data to evaluate the potential impact of missing data on our results. After running the binary logistic regression once more, we found consistent results with our initial models and equations. Moreover, these models continued to fit the data adequately.

Given our large sample size, it appears that the missing data didn’t impact our models negatively. Thus, our initial concerns regarding missing data exceeding the 20% threshold in some models were assuaged.

## Results

In our study, a total of 12 models were formulated across three age groups, each analyzed separately for men and women. The results of these models are detailed in two tables:

Table 4: This table displays results from the six models that encompass the categorical variable related to the ‘number of active voluntary memberships’. These models are designated as A + [age group] + [gender].

**Table 2:** Socioeconomic-Demographic Explanatory Variables.

| No. | Explanatory Variable | Description | Percentage of Dummy=1 |
| --- | --- | --- | --- |
| 1 | Marital Status: Married | Dummy set to 1 if individual is married; 0 if unmarried | 63.5% |
| 2 | Religious Person: Religious | Dummy set to 1 if individual is religious; 0 if not religious or an atheist | 61.5% |
| 3 | Employment Status: Paid Employment | Dummy set to 1 if individual has paid employment; 0 otherwise | 59.5% |
| 4 | Education: University | Dummy set to 1 if individual holds a university-level degree; 0 otherwise | 25.0% |
| 5 | Income: High Income | Dummy set to 1 if individual falls in high-income bracket; 0 for low income | 37.0% |
| 6 | Life Satisfaction: Satisfied | Dummy set to 1 if individual is satisfied with life; 0 if dissatisfied | 76.1% |
| 7 | Area of Living: Rural | Dummy set to 1 if individual resides in a rural area; 0 for urban area | 31.3% |
| 8 | Children | Dummy set to 1 if individual has children; 0 if childless | 70.5% |
| 9 | Political Regime: Free | Dummy set to 1 if individual lives in a country with a free political regime; 0 for 'not-free' political regimes | 38.7% |

**Table 3:** Binary Variables Pertaining to Voluntary Membership.

| Variable | Description |
| --- | --- |
| V1: Sport or Recreational Organization | Dummy set to 1 if active member; 0 otherwise |
| V2: Professional Association | Dummy set to 1 if active member; 0 otherwise |
| V3: Art, Music, or Educational Organization | Dummy set to 1 if active member; 0 otherwise |
| V4: Humanitarian or Charitable Organization | Dummy set to 1 if active member; 0 otherwise |
| V5: Church or Religious Organization | Dummy set to 1 if active member; 0 otherwise |
| V6: Labor Union | Dummy set to 1 if active member; 0 otherwise |
| V7: Political Party | Dummy set to 1 if active member; 0 otherwise |
| V8: Environmental Organization | Dummy set to 1 if active member; 0 otherwise |
| V9: Consumer Organization | Dummy set to 1 if active member; 0 otherwise |
| V10: Self-help Group or Mutual Aid Group | Dummy set to 1 if active member; 0 otherwise |
| V11: Women's Group | Dummy set to 1 if active member; 0 otherwise |
| V12: Other Organization | Dummy set to 1 if active member; 0 otherwise |

**Table 4:**
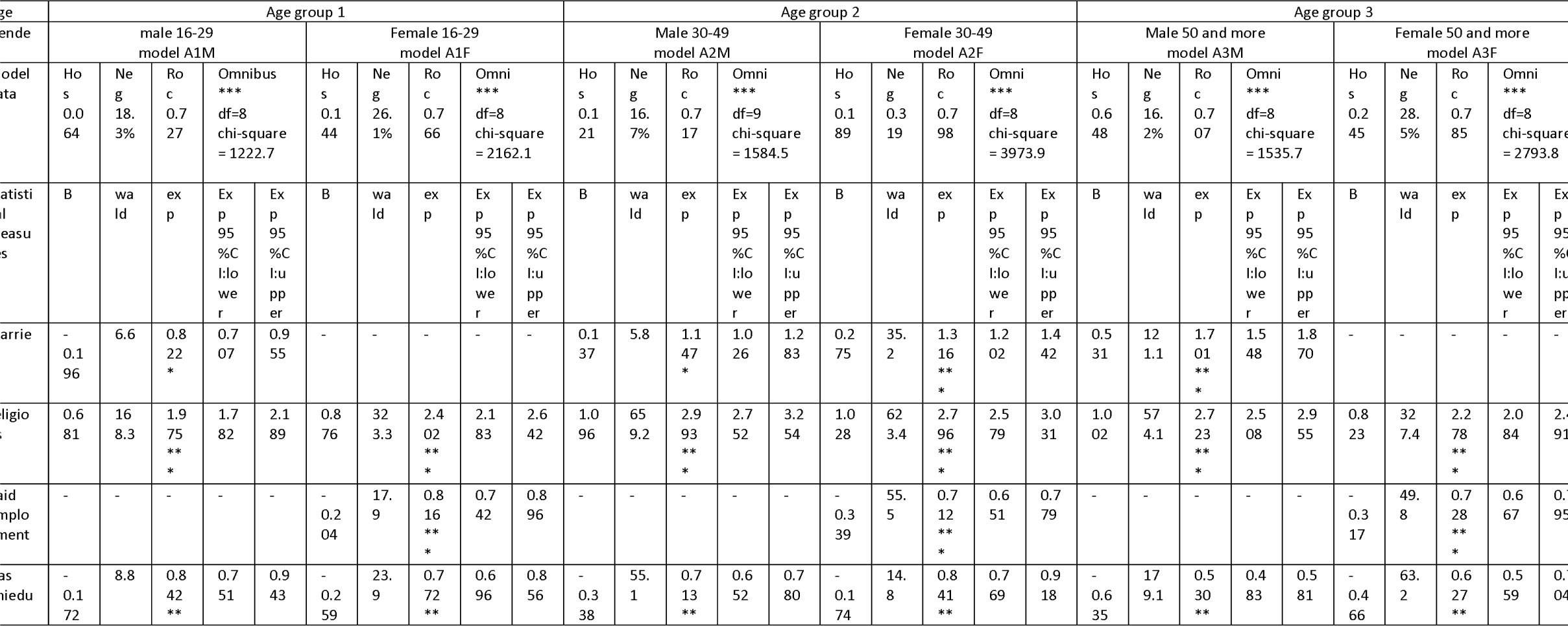
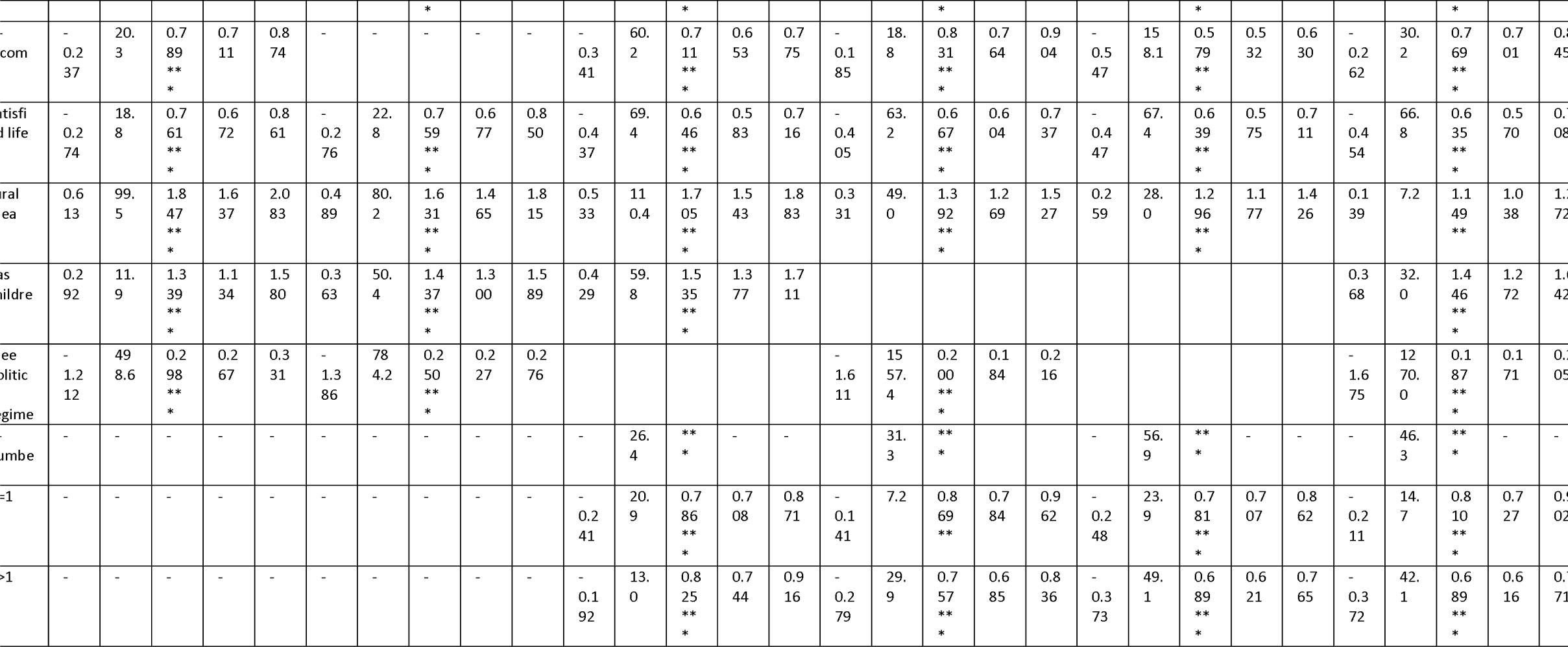
Group A Models Containing V-number Variable.

Table 5: This table showcases the Group B Models that incorporate variables related to the ‘type of voluntary organization’. Importantly, only respondents with active participation in either 0 or 1 voluntary organization were included. Models in this table are labeled as B + [age group] + [gender].

**Table 5:**
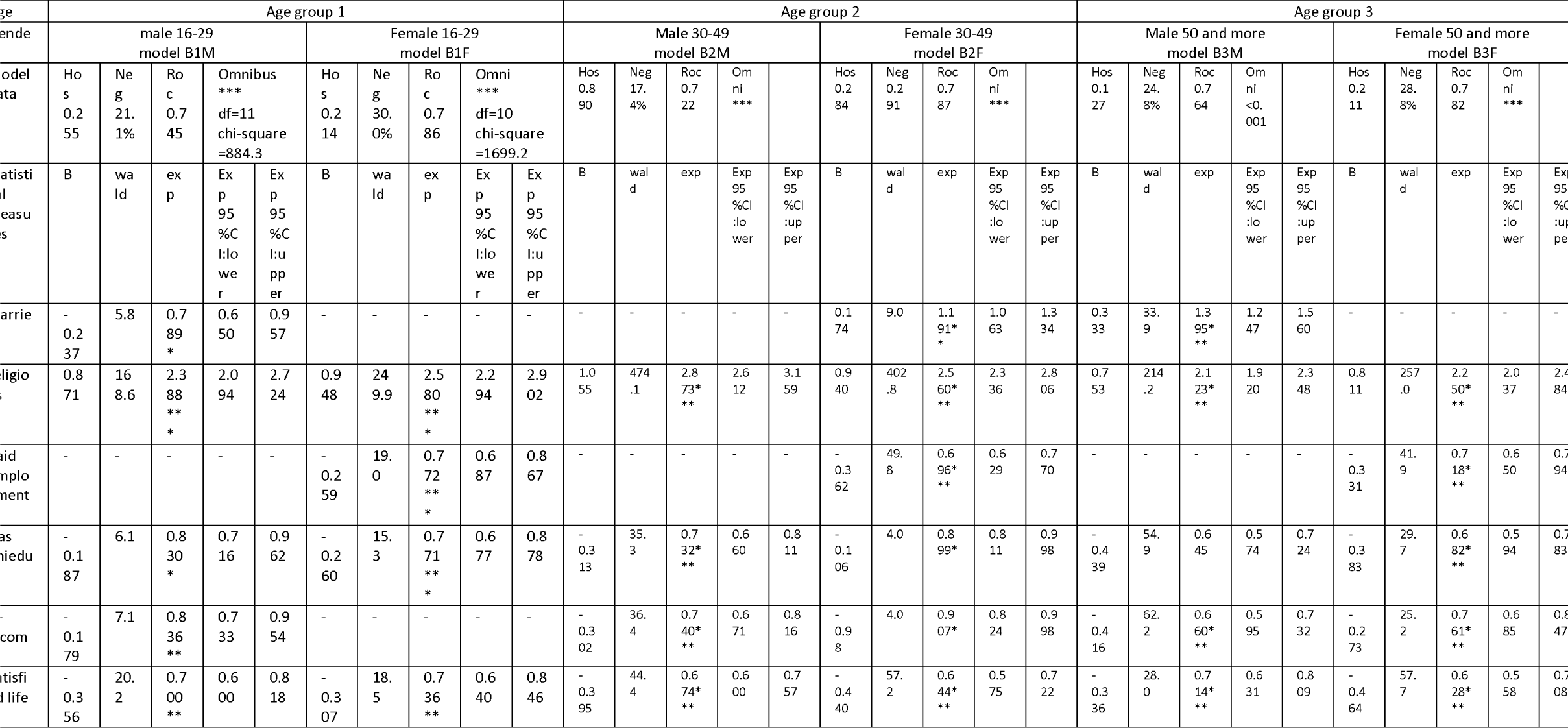
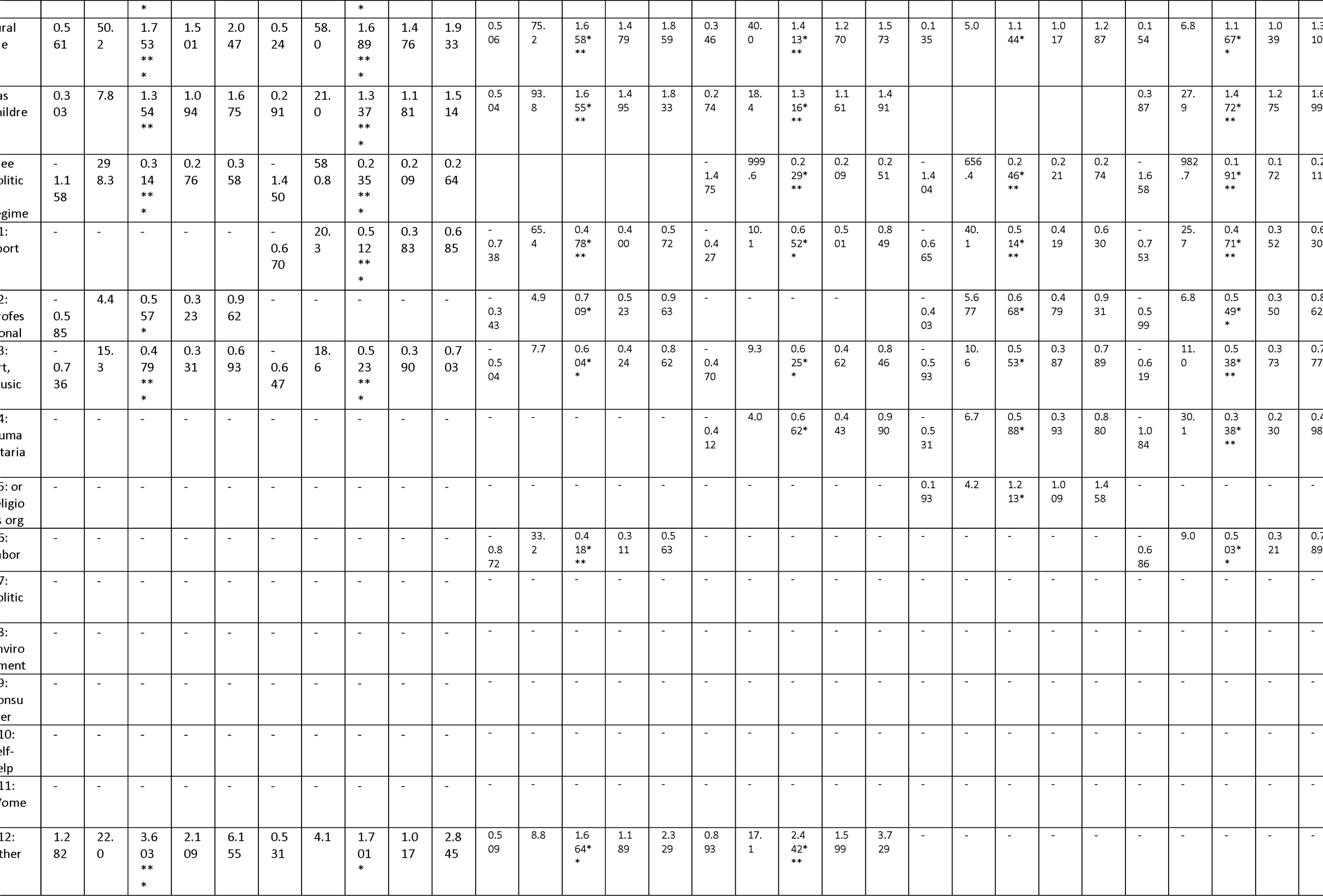
Group B Models with Voluntary Organization Types.

### Model Fitness

All formulated models demonstrated a robust fit to the data:

Omnibus tests consistently exhibited statistical significance.

Hosmer-Lemeshow tests remained non-significant across all models, indicating a good fit.

The Area Under the Curve (AUC) of the Receiver Operating Characteristic (ROC) consistently surpassed 0.7, a threshold generally accepted as indicating satisfactory classifier performance (Lu et al., 2011).

In most models, the Nagelkerke R^2 value exceeded 24%, and no model displayed a value below 16%.

### Variable Representation in Tables

Variables not intended for a specific model (like explanatory variables E8 and E9, which were included only if they did not adversely affect model fit) are denoted with ‘N/A’.

Variables initially planned for inclusion (like explanatory variables E1 through E7 and voluntary variables) but were later excluded due to a lack of statistical significance are marked with ‘-’.

### Summary of Results by Explanatory Variables (Derived from Group A Models)

#### Marital Status (E1)

Men: Marital status correlates with homonegativity across all age groups. Marriage decreases the odds of homonegativity by 17.8% in age group 1. However, it increases the odds by 14.7% in age group 2 and 70.1% in age group 3.

Women: Only in age group 2 does marital status increase the odds of homonegativity by 31.6%.

#### Religiousness (E2)

Religiousness is a potent predictor of homonegativity, increasing odds by 2-3 times. The association is least in men from age group 1 (1.975) and most pronounced in age group 2 (2.993).

#### Employment (E3)

Men: No discernible association.

Women: Employment reduces the odds of homonegativity by 18.4% in age group 1 and around 28% in age groups 2 and 3.

#### University Education (E4)

Having a university education reduces the odds of homonegativity across age groups, ranging from 15.8% to 47%. The decline is most marked in age group 3 with men at 47% and women at 37.3%.

#### Income (E5)

Having a high income decrease the odds of homonegativity across all groups, ranging from 16.9% to 42.1%. The sole exception is women in age group 1, where there’s no association.

#### Life Satisfaction (E6)

Satisfaction with life reduces the odds of homonegativity across the board, with a range from 23.9% to 46.5%.

#### Rural Area (E7)

Residing in rural areas increases the odds of homonegativity across all groups, with a variance from 14.9% to 84.7%.

#### Children (E8)

Having children boosts the odds of homonegativity in applicable age groups, ranging from 33.9% to 53.5%.

#### Free Political Regime (E9)

This variable was included in 4 out of 6 models, and it indicates a strong reduction in odds of homonegativity, ranging from 70.2% to 81.3%. When present in a model, it emerges as the strongest predictor of homonegativity compared to other variables.

### V-Number Variable

For age groups 2 and 3, an increase in the number of active memberships in voluntary organizations correlated with a decrease in homonegativity odds, especially pronounced when V > 1 (compared to V = 0).

Group B consists of six models tailored for individuals who are active members in either 0 or 1 voluntary organization. These models, named as B+age group+gender, aim to analyze the relationship between active membership in specific types of voluntary organizations and homonegativity. The findings are detailed in the following table (Table 5):

For each age-gender group, we highlight the key associations derived from Group B Models: Men, Age Group 1:

Active membership in V2 (Professional) and V3 (Art) decreases the odds of homonegativity by 41.5% and 26.4%, respectively. Conversely, V12 (Other) increases these odds by 3.6 times.

### Women, Age Group 1

V1 (Sport) and V3 (Art) memberships reduce the odds by 48.8% and 47.7% respectively. V12 (Other) boosts the odds by 70.1%.

### Men, Age Group 2

Memberships in V1, V2, V3, and V6 (Labor Union) diminish the odds by 52.2%, 29.1%, 39.6%, and 58.2% respectively. On the flip side, V12 (Other) membership raises the odds by 66.4%.

### Women, Age Group 2

Participations in V1, V3, and V4 (Humanitarian) result in odds reductions of 34.8%, 37.5%, and 33.8%, respectively. However, V12 (Other) pushes the odds up by a factor of 2.4.

### Men, Age Group 3

Engaging in V1, V2, V3, and V4 decreases the odds by 48.6%, 33.2%, 44.7%, and 41.2% respectively. In contrast, V5 (Religious Organizations) increases the odds by 21.3%.

### Women, Age Group 3

Involvement in V1, V2, V3, V4, and V6 reduces the odds by 52.9%, 45.1%, 46.2%, 66.2%, and 49.7% respectively. Notably, this is the only group where no voluntary activity, including V12, increases the odds of homonegativity.

## Discussion

The cautionary approach to data interpretation in our study arises from the variability in the inclusion of explanatory variables across our models. Notably, variables E8 (children) and E9 (free political regime) were integrated only when their presence didn’t jeopardize the model’s fit to the data. Hence, when aiming to compare variables across different models, it’s essential that both models exhibit a consistent inclusion pattern, particularly concerning E8 and E9. However, it’s worth noting that other variables, E1 to E7, found their way into all models as long as they demonstrated statistical significance.

To elucidate, within the framework of Group A models, a direct comparison is viable solely between men and women in age group 1, i.e., A1M and A1F. Both these models encapsulate E8 and E9 variables while excluding the V-number variable. Transitioning to Group B models, a similar logic applies: the data from females spanning age groups 1, 2, and 3 (i.e., B1F, B2F, B3F) and from males in age group 1 (B1M) can be juxtaposed directly, given their consistent incorporation of both E8 and E9.

When we analyze the data for men and women in age group 1, referencing Group A models (A1M and A1F), several key insights emerge:

Marital Status: Being married or cohabiting doesn’t impact homonegativity levels among women. However, for men, there’s a notable association, revealing a 17.8% decrease in homonegativity odds, albeit with a fairly broad 95% confidence interval (CI).

Income Levels: Higher income levels in men are correlated with a 21.1% reduction in homonegativity odds. Such an association isn’t present among the women of this age group.

Religiosity: Women in this age group exhibit a more robust link between religiousness and homonegativity, with an odds ratio of 2.40, as opposed to men, who have an odds ratio of 1.97.

Employment: There’s no discernible link between employment and homonegativity in men. In contrast, women in employment have an 18.4% reduced likelihood of exhibiting homonegative tendencies.

Educational Attainment: Possessing a university-level education correlates with decreased homonegativity for both genders, by 15.8% for men and 22.8% for women.

Life Satisfaction: For both men and women in this age bracket, greater life satisfaction relates to a roughly 24% reduction in homonegativity odds.

Residential Setting: Residing in rural areas is associated with increased homonegativity for both genders – a substantial 84.7% increase for men and a 63.1% increase for women.

Parental Status: For men, being a parent augments the odds of homonegativity by 33.9%, whereas for women, the increment is slightly higher at 43.7%.

Political Landscape: Inhabiting a nation with a free political regime emerges as a significant factor. It drastically diminishes the likelihood of homonegativity by 70.2% for men and 75% for women. Intriguingly, this variable exerts the strongest influence on homonegativity compared to all others; when you invert its odds ratio, its impact even surpasses that of the religiosity variable.

In assessing the associations of homonegativity with specific types of voluntary organizations through Group B models, several key relationships were identified:

### For Men in Age Group 1

Active membership in a professional voluntary organization (V2) was linked to a 44.3% reduction in homonegativity odds, though this finding had a wide 95% confidence interval. Membership in art-music-educational organizations (V3) led to an even more significant 52.1% drop in homonegativity odds.

### For Women in Age Group 1

Active participation in a voluntary sports organization (V1) was associated with a 48.8% decline in the odds of homonegativity. Engaging with art-music-educational organizations (V3) mirrored this trend, with a 47.7% reduction.

Conversely, affiliation with ‘other’ voluntary organizations (V12) escalated homonegativity odds. Men saw this increase threefold, while women faced a 70.1% rise. It’s noteworthy that the specific nature and objectives of these ‘other’ organizations remain unidentified.

When we broadened our analysis across women in age groups 1, 2, and 3 (B1F, B2F, B3F) using Group B models, the following patterns were discernible:

Membership in voluntary sports organizations diminished homonegativity odds by 48.8%, 34.8%, and 52.9% across the three age groups, respectively. Affiliation with art-music-educational organizations similarly reduced odds by 47.7%, 37.5%, and 46.2%. Professional organization membership showed a clear reduction in homonegativity only for women in the third age group, by 45.1%. Engagement with humanitarian organizations cut back homonegativity odds in age groups 2 and 3 by 37.8% and 66.2% respectively. Active participation in a Labor Union led to a 49.7% reduction in homonegativity for women in the third age group. The enigmatic V12 (’other’ voluntary organizations) augmented the odds of homonegativity by 66.4% and more than double for women in the first and second age groups, respectively. Men aged 30-50 saw a significant 58.2% reduction in homonegativity when affiliated with a Labor Union. However, no associations were spotted for men in age groups 1 and 3.

Lastly, several voluntary organizations, encompassing political (V7), environmental (V8), consumer (V9), self-help (V10), and women-oriented ones (V11), showed no discernible relationship with homonegativity in any of the models under scrutiny.

Expanding on the findings from Group A models, individuals in age groups 2 and 3 who maintain an active membership in voluntary organizations exhibit a decrease in homonegativity odds ranging from 14-22%. Interestingly, holding memberships across multiple organizations amplifies this effect: those in age groups 2 and 3 show a 18-31% reduction in homonegativity odds, indicating that diverse engagements might foster more inclusive mindsets.

While these results undeniably highlight the negative correlation between active membership in voluntary organizations and homonegativity, it’s crucial to avoid conflating correlation with causation. The data doesn’t definitively argue that voluntary participation directly mitigates homonegativity. However, considering the evidence, it remains a viable theory.

Supporting this potential causative link, literature points out that volunteering cultivates a richer environment for interactions, stimulating dialogues that create safe havens for self-expression. Such environments foster mutual understanding, and tolerance (Rusu, 2016). Moreover, active participation in voluntary organizations has been linked to the development of generalized trust, mutual respect and toleration (Wilson & Musick, 1999). Given these premises, it stands to reason that if voluntary engagements bolster general tolerance, they might very well also influence a more accepting stance towards homosexuality. Consequently, it seems plausible that the active involvement in such organizations encourages greater acceptance of minorities, including the LGBTQ+ community. This underlying correlation could indeed hint at a causative relationship, making the findings of this study all the more significant.

## Conclusion

Demographic and socio-economic variables play a discernible role in shaping attitudes towards homonegativity. However, their impact varies across distinct gender-age groups. Notably, a higher life satisfaction tends to diminish homonegative sentiments. Furthermore, the political freedoms characterizing a respondent’s home country emerge as the most potent predictor, inversely related to homonegativity.

Our analysis has further illuminated the relationship between active participation in various voluntary organizations and homonegativity. Specifically, engagements with sport or recreational organizations, professional associations, art, music, or educational organizations, as well as humanitarian or charitable entities, often correlate with reduced homonegative tendencies among certain gender-age groups. Notably, every gender-age group highlighted in the study had at least two voluntary organizations where active participation was linked to decreased homonegativity.

However, it’s also essential to mention the presence of certain voluntary organizations where membership seemed to amplify homonegative views. The specifics of these organizations remain unidentified in our current data.

## Data Availability

All data produced are available online at
https://www.worldvaluessurvey.org/wvs.jsp
any additional data(data analysis methods) is available upon request.

## Declarations

### Ethical Approval and Consent to Participate

We utilized secondary data from the World Values Survey and obtained permission via email to use this data. The study received IRB approval from Chulalongkorn University (IRB Number: 0683/66).

### Availability of Data and Materials

Data can be accessed on the World Values Survey website: [https://www.worldvaluessurvey.org/wvs.jsp](https://www.worldvaluessurvey.org/wvs.jsp)

### Competing Interests

Not applicable.

### Funding

Chulalongkorn University faculty of Medicine

## Acknowledgements

Not applicable.

## Notes

### Competing Interest Statement

The authors have declared no competing interest.

### Author Declarations

We utilized secondary data from the World Values Survey (https://www.worldvaluessurvey.org/wvs.jsp) and obtained permission via email to use this data. The study received IRB approval from Chulalongkorn University (IRB Number:0683/66).

